# Mild Behavioural Impairment-Apathy and Alzheimer’s Disease Plasma Phosphorylated Tau Biomarker Levels

**DOI:** 10.64898/2026.02.25.26347102

**Authors:** Daniella Vellone, Rebeca Leon, Zahra Goodarzi, Nils D. Forkert, Eric E. Smith, Zahinoor Ismail

## Abstract

**Background:** Mild behavioural impairment (MBI), characterized by later-life emergence of persistent neuropsychiatric symptoms (NPS), is an early clinical indicator of dementia risk. MBI as a global construct has been associated with Alzheimer disease (AD) pathology; studies have also explored MBI domains. Prior work has linked MBI-apathy to cerebrospinal fluid (CSF) biomarkers of AD, but whether similar associations are detectable using plasma-based biomarkers such as phosphorylated tau (p-tau) is unknown. Establishing such relationships is critical, as plasma biomarkers are more accessible than CSF.

**Objective:** To explore cross-sectional and longitudinal associations between MBI-apathy and plasma p-tau_181_ using Alzheimer’s Disease Neuroimaging Initiative data.

**Methods:** Older adults with normal cognition or mild cognitive impairment were categorized as MBI-apathy (n=69), non-MBI NPS (n=112), and no-NPS (n=215) based on Neuropsychiatric Inventory scores and symptom persistence over one year. Linear regression modelled cross-sectional associations between NPS group and plasma p-tau_181_ levels, adjusting for age, sex, education, apolipoprotein E4 status, and Mini-Mental State Examination score. Hierarchical linear mixed-effects modelling assessed associations over two and three years, including time-by-NPS group interactions.

**Results:** MBI-apathy was associated with significantly higher plasma p-tau_181_ levels at baseline (24.05% [6.06–45.08%]; adjusted *p*=0.014), and over two (26.46% [7.24–49.12%]; adjusted *p*=0.012) and three years (29.28% [10.17–51.72%]; adjusted *p*=0.004) compared to no-NPS. No significant associations were observed for non-MBI NPS.

**Conclusions:** MBI-apathy is associated with elevated plasma p-tau_181_ cross-sectionally and longitudinally. These findings support MBI-apathy as a potential proxy marker of tau pathology for early AD detection.

## Introduction

Alzheimer’s disease (AD) is a progressive neurodegenerative condition characterised by the accumulation of pathological tau and amyloid-β proteins (Aβ).^1^ Biomarker-based criteria are now central to the diagnosis and staging of AD, shifting the field towards incorporating AD-specific biology into what was primarily a clinical definition of the disease.^1^ In the Alzheimer’s Association 2024 revised AD criteria,^1^ the biological definition of AD is grounded in a two-tiered biomarker classification system for diagnosis and staging. Core 1 biomarkers are essential for diagnosis and include measures of amyloid and tau. Specifically, these markers include A for Aβ (*i.e.,* cerebrospinal fluid [CSF] and blood plasma Aβ_42_, as well as amyloid positron emission tomography [PET]) and T1 for phosphorylated tau [p-tau], (*i.e.,* CSF and blood plasma p-tau_181/217/231_). In contrast, Core 2 biomarkers are not required for diagnosis but can support staging and further biological characterization. Core 2 biomarkers reflect later-stage tau pathology (T2), including microtubule-binding region tau_243_, other phosphorylated tau forms (*e.g.,* p-tau_205_), non-phosphorylated mid-region tau fragments, and tau PET imaging.

The earliest detectable stage of AD pathophysiology is marked by amyloid accumulation.^2^ As soluble Aβ is sequestered into plaques, CSF Aβ_42_ levels decline, and amyloid PET signal increases, often years before the onset of clinical symptoms.^3–8^ Following amyloid plaque deposition, tau becomes aberrantly phosphorylated at multiple mid-region residues, reflecting downstream effects of Aβ-driven kinase dysregulation and glial activation.^1, 3–5, 9^ Distinct phospho-epitopes, including threonine 181 (p-tau_181_) and threonine 217 (p-tau_217_), show overlapping but temporally distinct trajectories.^3–5^ P-tau_217_ rises earliest and more steeply with emerging Aβ pathology, whereas p-tau_181_ increases slightly later and plateaus more gradually during the prodromal phase.^10^ Phosphorylation at threonine 181 likely represents an early but downstream response to amyloid plaque formation, preceding the development of insoluble tau aggregates detectable by tau PET.^1, 5, 11^ Consequently, phosphorylated tau species function as intermediate markers linking amyloid deposition to later neurodegenerative changes. Although p-tau_217_ has demonstrated superior sensitivity and specificity for detecting early AD pathology,^3, 11–17^ p-tau_181_ remains a valuable biomarker for longitudinal designs, as its slower, more stable trajectory provides insight into disease progression across preclinical and prodromal stages. The reliable detectability of p-tau_181_ in plasma enhances scalability for early identification of AD pathophysiology, especially in settings where access to neuroimaging or CSF collection is limited. The integration of fluid biomarkers into diagnostic workflows represents a pivotal shift toward earlier and more efficient identification of individuals at risk.

Although cognitive symptoms have traditionally provided the impetus for AD assessment and work up, neuropsychiatric symptoms (NPS) are recognized as early manifestations of the disease. In particular, mild behavioural impairment (MBI) is a validated framework for dementia detection and prognostication that leverages risk associated with later-life emergent and persistent NPS. Multiple studies have demonstrated that MBI is associated with incident cognitive decline and dementia.^18–22^ MBI comprises five behavioural domains, including apathy, affective dysregulation, impulse dyscontrol, social inappropriateness, and psychosis, some of which have also been studied longitudinally for risk of incident cognitive decline and dementia.^23–26^ Apathy in neurocognitive disorders is defined as diminished interest, initiative, and/or emotional reactivity,^27–30^ and is particularly salient given its strong associations with tau pathology and progression to AD dementia.^24, 31^ In earlier work, we demonstrated that MBI-apathy was associated with greater risk of incident dementia, even in the absence of cognitive impairment.^24^ More recently, we found that MBI-apathy is associated with greater CSF tau pathology.^31^ Few studies, however, have examined whether these associations extend to plasma-based biomarkers in non-dementia populations.^32, 33^ Clarifying the relationship between MBI-apathy and plasma biomarkers is important, as these biomarkers are less invasive and more scalable than CSF or imaging, making them well suited for real-world clinical implementation. As MBI-apathy is a clinically observable and easily screenable behavioural syndrome, establishing alignment with plasma tau pathology could provide a pragmatic avenue for early risk stratification in pre-dementia populations.

Accordingly, the objective of this study was to investigate associations between MBI-apathy and plasma p-tau_181_ in individuals who are cognitively normal (CN) or have mild cognitive impairment (MCI). We hypothesized that MBI-apathy is associated with elevated plasma p-tau_181_ levels at baseline and that this association persists longitudinally over two- and three-year follow-up periods.

## Materials and Methods

### Study Cohort & Data Source

This study utilized data from the Alzheimer’s Disease Neuroimaging Initiative (ADNI; https://adni.loni.usc.edu/), a large, multicentre, longitudinal study launched in 2003 under the leadership of Principal Investigator Michael W. Weiner, MD. Supported by both public and private funding, ADNI was established to develop standardized clinical, imaging, genetic, and biomarker methodologies for assessing individuals across the cognitive spectrum, from normal cognition to mild AD. The dataset includes comprehensive demographic, diagnostic, neurological, genetic, and neuropathological assessments collected at six- to twelve-month intervals. ADNI adheres to Good Clinical Practice guidelines, the ethical principles of the Declaration of Helsinki, and U.S. regulatory requirements (21 CFR Part 50: Protection of Human Subjects and Part 56: Institutional Review Boards). The study protocol was approved by the Institutional Review Boards at all participating sites. All participants provided written informed consent for participation and publication prior to the initiation of any study procedures.

Participant privacy was protected through the use of coded research identifiers; no personally identifying information was linked to the data used in this analysis. Data and samples were collected from September 2005 to September 2023. The available ADNI data used in this work were accessed and downloaded on March 11^th^, 2024.

### Participant Inclusion Criteria & Classification

The study flow for primary data analysis is shown in Figure 1. Cognitive diagnoses (CN or MCI) were based on the Mini Mental State Examination (MMSE) and Clinical Dementia Rating (CDR). CN participants had an MMSE score ≥ 24 and a CDR score of 0. Participants with MCI had an MMSE score ≥ 24, a CDR score of 0.5, memory complaints, and objective memory impairment based on education-adjusted scores from the Wechsler Memory Scale Logical Memory II, with preserved activities of daily living.

**Figure 1.**
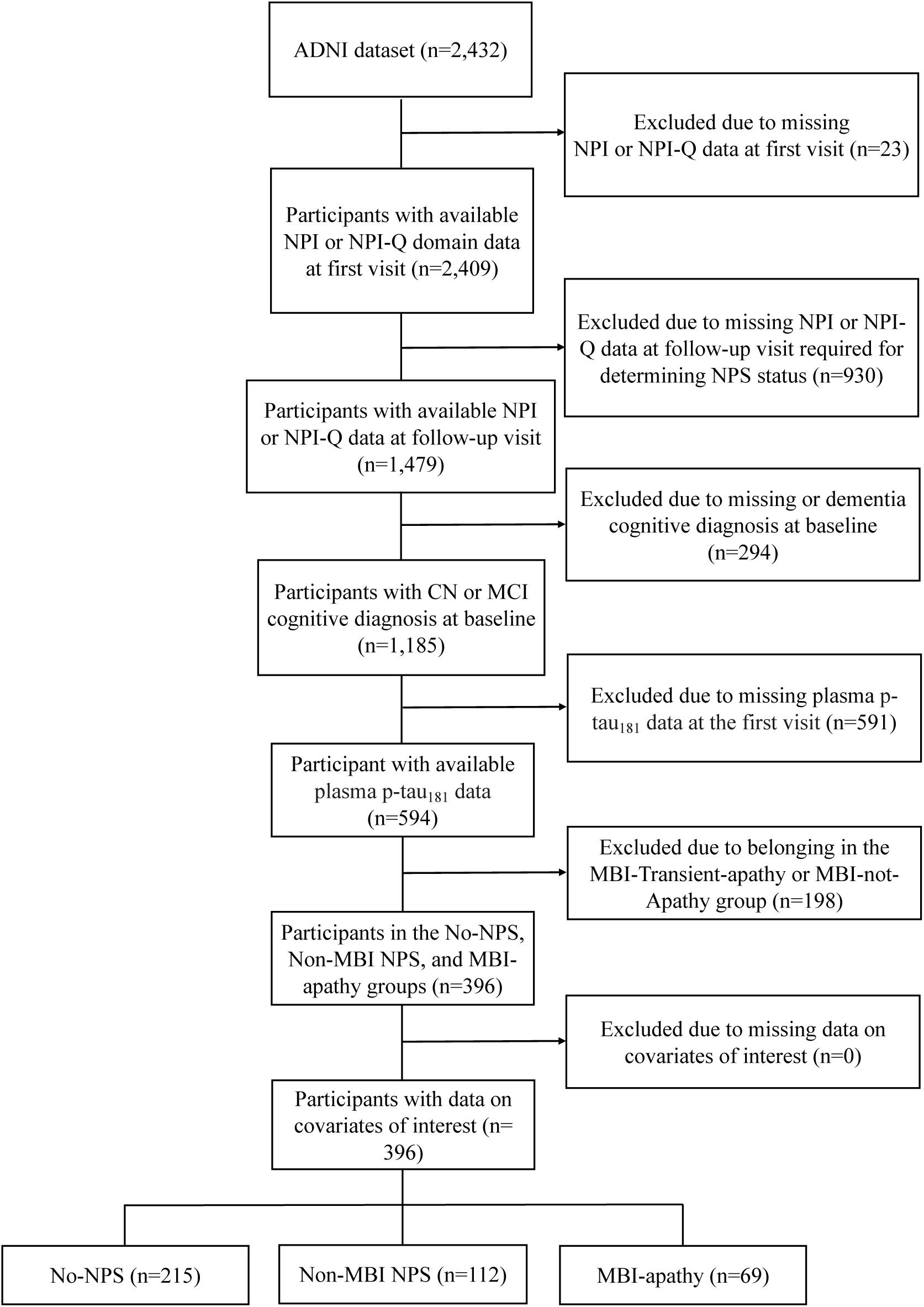
Flowchart of participants from ADNI included for analysis. ADNI: Alzheimer’s Neuroimaging Initiative; CN: Cognitively Normal; MBI: Mild Behavioural Impairment; MCI: Mild Cognitive Impairment; NPI: Neuropsychiatric Inventory; NPI-Q: Neuropsychiatric Inventory Questionnaire; NPS: Neuropsychiatric Symptoms

The classification of participant groups in this study follows a previously established methodology.^31^ Participants were included if they had complete Neuropsychiatric Inventory (NPI)^34^ (from ADNI-2, 3) or Neuropsychiatric Inventory Questionnaire (NPI-Q)^35^ (from ADNI-1, GO, 2, 3) domain scores necessary for determining NPS status and at least two study visits within their first year to assess symptom persistence.

As previously described,^31^ MBI status was determined by transforming NPI and NPI-Q scores into MBI domain scores using a published algorithm.^36^ MBI-apathy was operationalized based on the NPI and NPI-Q apathy domain scores. The presence of apathy was defined as an apathy domain score > 0, with symptom persistence requiring identification in at least two of three visits within the first year (*i.e.,* 0 and 6 months, 0 and 12 months, 6 and 12 months, or all three time points). Individuals were included in the MBI-apathy group irrespective of concurrent NPS in other MBI domains.

Participants who exhibited NPS in any of the five MBI domains (apathy, affective dysregulation, impulse dyscontrol, social inappropriateness, psychosis) at only one of the three visits within the first year were classified as the non-MBI NPS group, as they did not meet the MBI symptom persistence criterion. The no-NPS group included participants without any NPS (MBI total score = 0) at all first-year visits.

### Biomarker Analysis: Plasma p-tau181

Participants were required to have baseline plasma p-tau_181_ measurements for inclusion in this analysis. ADNI plasma p-tau_181_ concentrations (ng/L) were collected annually and analyzed in a blinded manner between May and December 2019 using the Single Molecule Array (Simoa) HD-1 platform (Quanterix, Billerica, MA, USA) in the Clinical Neurochemistry Laboratory at the University of Gothenburg, Sweden, according to established protocols.^37^ The assay employs Tau12 and AT270 monoclonal antibodies to detect N-terminal to mid-domain forms of p-tau_181_. Further details on analytical procedures and assay validation are available elsewhere.^37^

### Statistical Analyses

Baseline demographic characteristics comprised age, sex, and years of education. Clinical data encompassed NPI/NPI-Q and MMSE scores, and biomarker measures included apolipoprotein E4 (APOE4) carrier status and plasma p-tau_181_ levels. Comparisons between the MBI-apathy and non-MBI NPS group versus the no-NPS group were performed using χ^2^ tests for categorical variables and one-way ANOVAs for continuous variables.

Both cross-sectional and longitudinal associations of NPS status and plasma p-tau_181_ were examined. For the cross-sectional analysis, a linear regression model was employed with NPS status as the predictor and plasma p-tau_181_ level as the continuous outcome variable. NPS status was categorized into three groups: MBI-apathy, non-MBI NPS, and no-NPS, with the no-NPS group serving as the reference. The model was adjusted for age, sex, years of education, APOE4 carrier status, and MMSE score. APOE4 carrier status was dichotomized, with individuals carrying one or more APOE4 alleles classified as APOE4 carriers and those without APOE4 alleles as non-carriers, with non-APOE4 carriers as the reference. Male was the reference group for sex.

To address skewness in plasma p-tau_181_ level, logarithmic transformations were applied. Additionally, values were Winsorized at the 5^th^ and 95^th^ percentiles to mitigate the influence of extreme outliers while preserving all participant data. The transformed distribution was then visually inspected with diagnostic plots (Residuals *vs.* Fitted, Normal Q-Q, Scale-Location, and Residuals *vs.* Leverage) to evaluate linearity, normality of errors, homoscedasticity, and the influence of potential outliers. The visual checks confirmed that assumptions required for regression analyses were adequately met.

Longitudinal associations between NPS status (predictor) and plasma p-tau_181_ levels (outcome variable) over two and three years were assessed using hierarchical linear mixed-effects (LME) models. Plasma p-tau_181_ levels were treated as repeated measures, whereas all other variables were assessed at baseline. To account for individual variability, participant ID was modeled with random intercepts and slopes. Fixed effects included NPS status, age, sex, years of education, APOE4 carrier status, and MMSE score. Additionally, an interaction term between NPS status and time in months was included to test whether biomarker trajectories differed across NPS groups relative to the no-NPS group.

All statistical analyses were performed using R v4.4.0. Linear models were implemented with the stats package, LME models with the lmer package, and group comparisons over time were examined using the emmeans package. To account for multiple testing and control the false discovery rate (FDR), we applied the Benjamini-Hochberg procedure to the *p*-values obtained from our analyses. Statistical significance was defined as an adjusted *p*-value < 0.05.

## Results

### Participant Demographics and Characteristics

The current study included 396 participants, with 215 in the no-NPS group, 112 in the non-MBI NPS group, and 69 in the MBI-apathy group. On average, participants were 72.5 years old, 49.5% female, and 41.4% CN. Compared to the no-NPS group, individuals with MBI-apathy were of similar age and education level, but were more likely to be male, APOE4 carriers, and have MCI (Table 1).

**Table 1.**
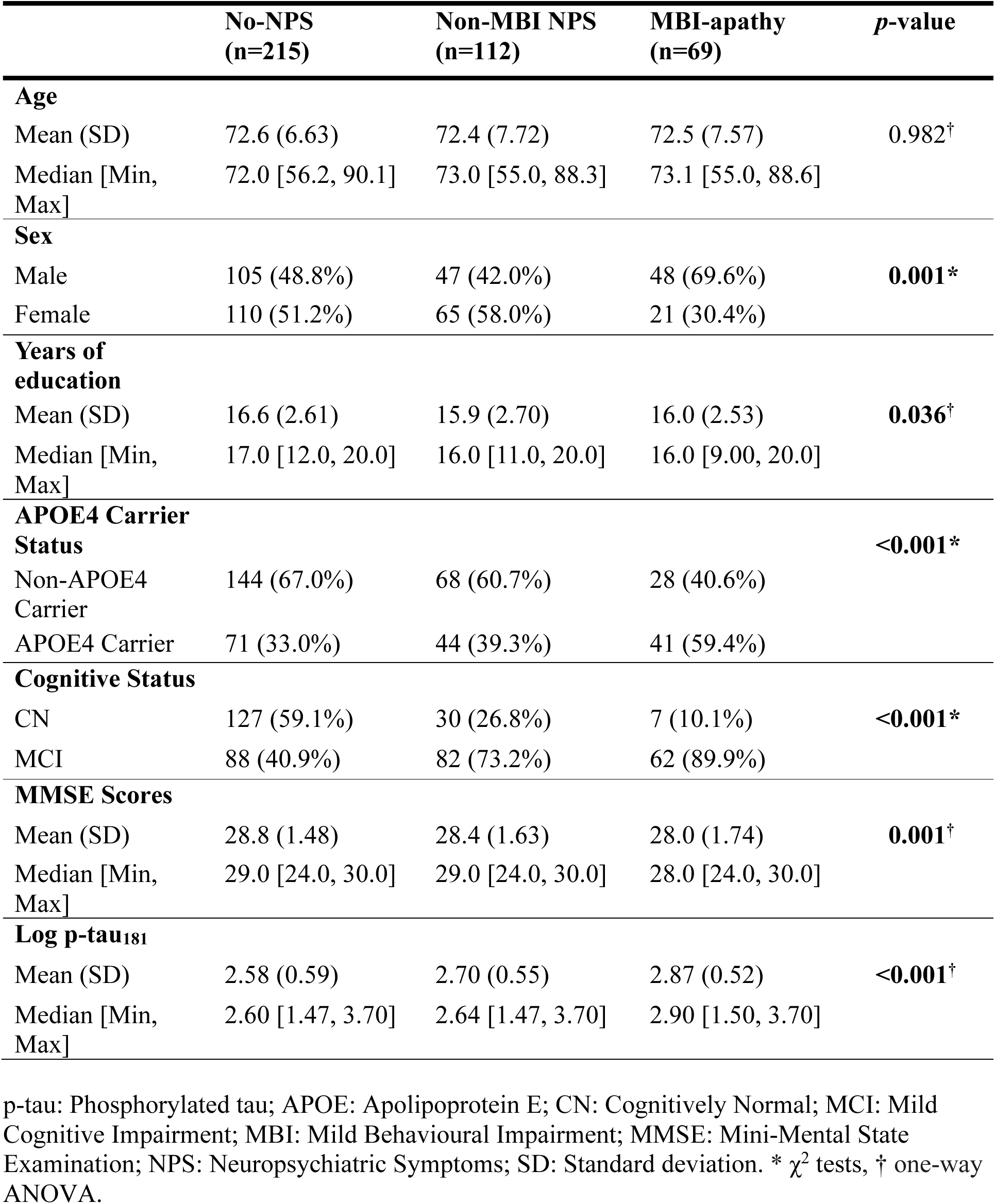
Baseline participant characteristics.

Plasma p-tau_181_ levels were significantly higher in the MBI-apathy group compared to the no-NPS group (*p*<0.001). A violin plot for baseline plasma p-tau_181_ level by NPS group is presented in Figure 2.

**Figure 2.**
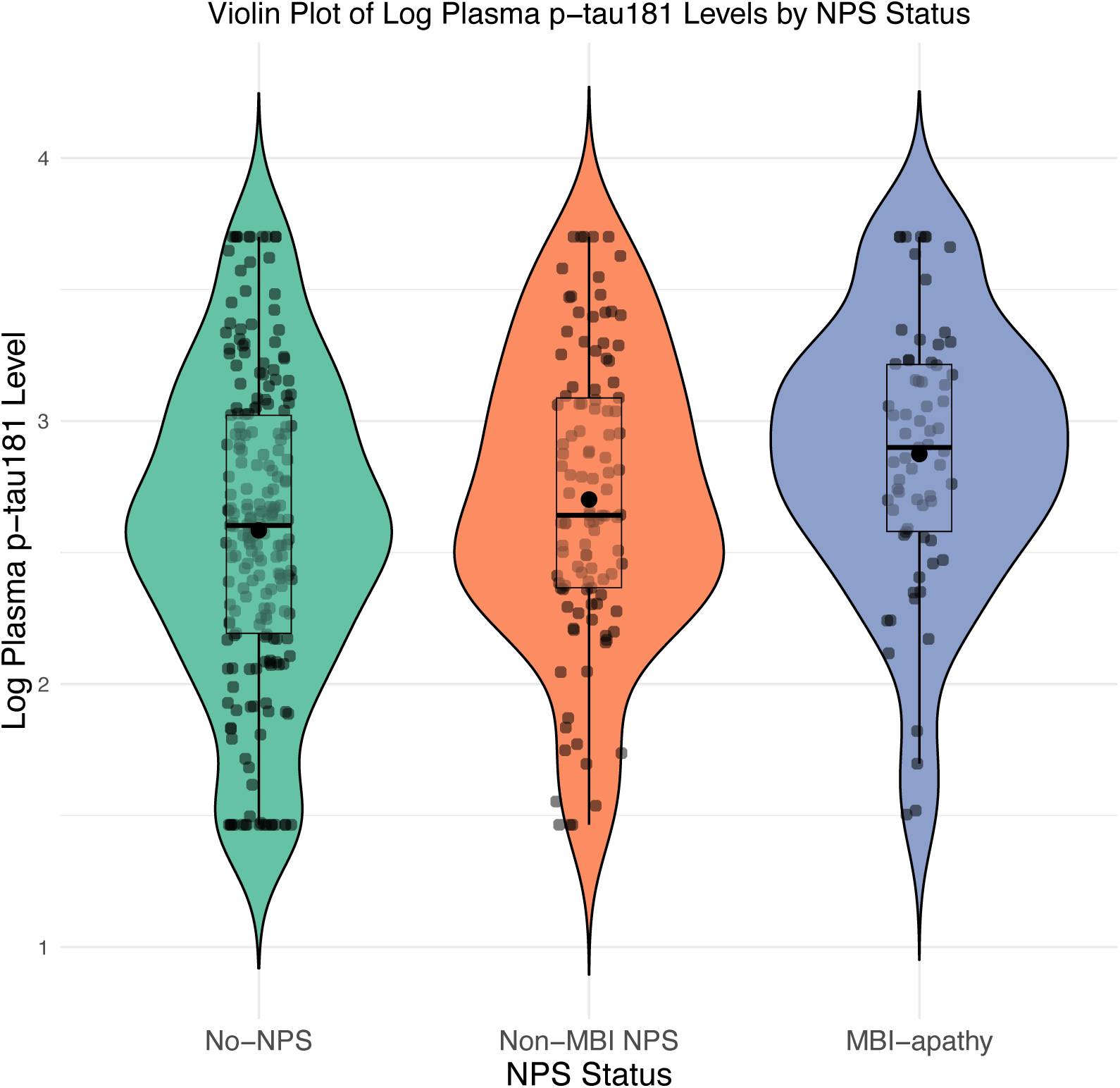
Violin plot showing the distribution of log plasma p-tau_181_ levels by NPS status, including jittered data points, boxplots for interquartile range and median, and mean values. P-tau: Phosphorylated tau; APOE: MBI: Mild Behavioural Impairment; NPS: Neuropsychiatric Symptoms.

### Cross-Sectional & Longitudinal Associations Between MBI-apathy and Plasma p-Tau181

The cross-sectional linear regression model revealed that MBI-apathy status was associated with 24.05% higher baseline plasma p-tau_181_ levels compared to the no-NPS group (percentage difference β [95%CI], 24.05% [6.06% – 45.08%]; FDR-adjusted *p*=0.014) (Table 2).

**Table 2.** Cross-sectional association between NPS group and plasma p-tau_181_ modelled using linear regression adjusted for covariates.

| Predictors | Percentage Difference $\beta$ | 95% CI | $p$-value | FDR-adjusted $p$-value |
| --- | --- | --- | --- | --- |
| Non-MBI NPS | 10.64% | -2.62% – 25.70% | 0.120 | 0.120 |
| MBI-apathy | 24.05% | 6.06% – 45.08% | **0.007** | **0.014** |
| Age | 1.57% | 0.76% – 2.38% | **<0.001** |  |
| Sex (Female) | -1.27% | -12.23% – 11.06% | 0.830 |  |
| Years of Education | 0.50% | -1.73% – 2.79% | 0.663 |  |
| APOE4 Carrier Status (APOE4 Carrier) | 23.69% | 9.84% – 39.28% | **<0.001** |  |
| MMSE Score | -2.65% | -6.18% – 1.02% | 0.154 |  |

Comparatively, non-MBI NPS status was not significantly associated with plasma p-tau_181_ levels (β=10.64% [-2.62% – 25.70%]; FDR-adjusted *p*=0.120) at baseline.

Longitudinal biomarker data for plasma p-tau_181_ were available for 328/396 participants at the two-year mark (Table 3 and Figure 3).

**Figure 3.**
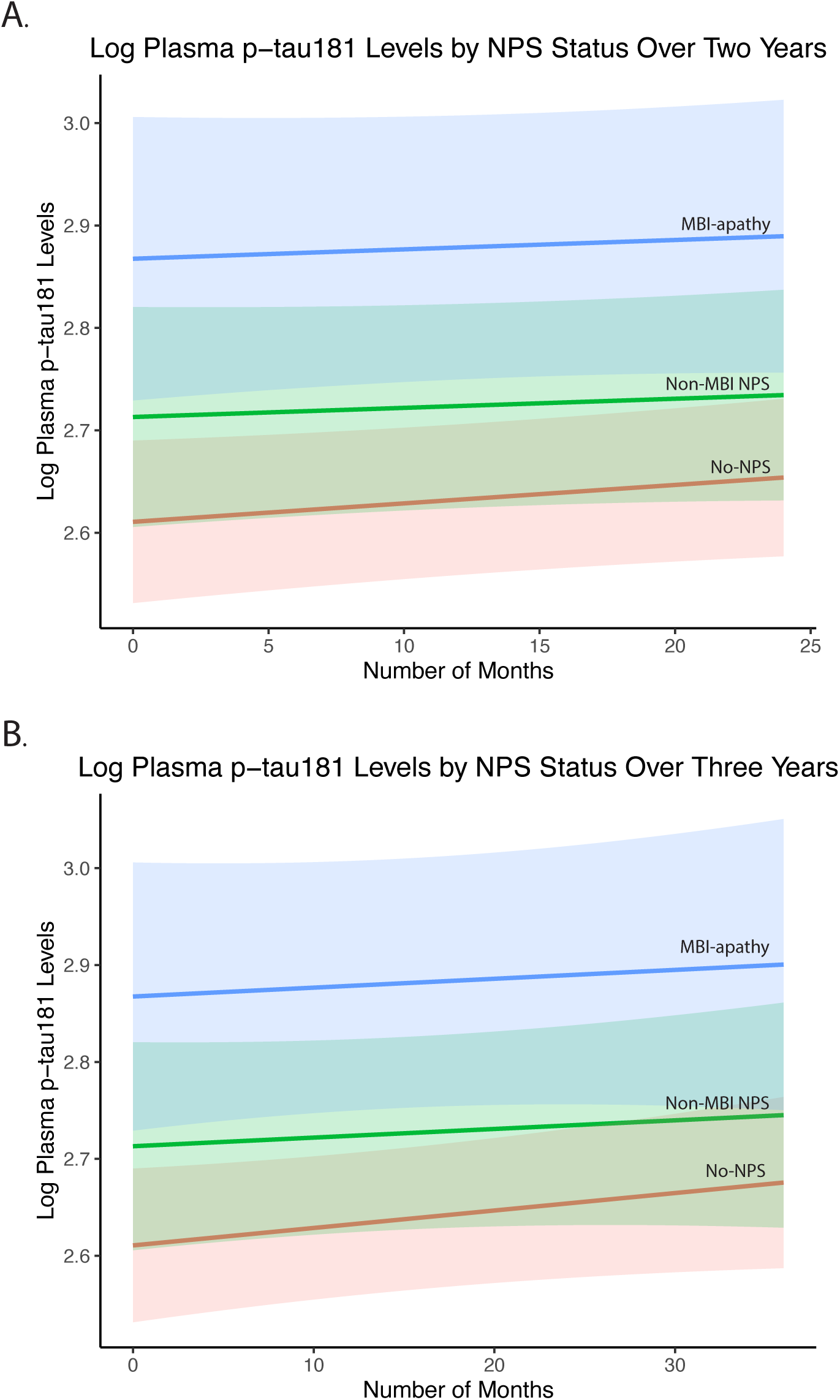
Longitudinal trajectories of plasma p-tau_181_ over time stratified by NPS status. Panels display estimated marginal mean trajectories with 95% confidence intervals for a. p-tau_181_ over two years and b. plasma p-tau_181_ over three years, stratified by NPS status: MBI-apathy, non-MBI NPS, and no-NPS (reference group). p-tau: Phosphorylated tau; MBI: Mild Behavioural Impairment; NPS: Neuropsychiatric Symptoms.

**Table 3.** Longitudinal association between NPS group and plasma p-tau_181_ over two years.

| Predictors | Percentage Difference $\beta$ | 95% CI | *p*-value | FDR-adjusted *p*-value |
| --- | --- | --- | --- | --- |
| Non-MBI NPS | 11.18% | -2.84% – 27.23% | 0.126 | 0.126 |
| MBI-apathy | 26.46% | 7.24% – 49.12% | **0.006** | **0.012** |
| Time (Months) | 0.15% | -0.14% – 0.43% | 0.318 |  |
| Age | 1.96% | 1.17% – 2.75% | **<0.001** |  |
| Sex (Female) | -5.77% | -15.92% – 5.60% | 0.310 |  |
| Years of Education | 0.12% | -2.04% – 2.33% | 0.914 |  |
| APOE4 Carrier Status (APOE4 Carrier) | 22.23% | 8.93% – 37.15% | **0.001** |  |
| MMSE Score | -2.53% | -5.96% – 1.03% | 0.165 |  |
| NPS status (Non-MBI NPS) × Time | -0.14% | -0.63% – 0.35% | 0.580 | |
| NPS status (MBI-apathy) × Time | 0.22% | -0.38% – 0.82% | 0.479 | |

**modelled using linear mixed effect models adjusted for covariates**

Beta coefficients for the NPS group variables (highlighted in grey) represent the estimated percent difference in p-tau_181_ compared to the no-NPS group over two years. Tests of interaction between NPS group and time were non-significant. This model is adjusted for age, sex, years of education, APOE4 carrier status, MMSE score, and an interaction term between NPS status and number of months. APOE: Apolipoprotein E; CI: Confidence interval; FDR: False Discovery Rate; MBI: Mild Behavioural Impairment; MCI: Mild Cognitive Impairment; MMSE: Mini-Mental State Examination; NPS: Neuropsychiatric Symptoms.

Participants missing plasma p-tau_181_ data at two years were older (74.63 [6.77] *vs*. 72.05 [7.10] years, *p* = 0.005) and had lower MMSE scores (28.03 [1.88] *vs.* 28.62 [1.51], *p* = 0.017) compared to those with data available at that timepoint. Sex distribution (Female: 52.94% *vs.* 48.78%, *p* = 0.623), years of education (15.79 ± 2.86 *vs.* 16.44 ± 2.58, *p* = 0.087), and APOE4 carriership (48.53% *vs.* 37.50%, *p* = 0.119) did not significantly differ between groups. On average, plasma p-tau_181_ levels did not significantly change over two years across the full sample (percentage change β [95%CI], 0.15% [-0.14% – 0.43%]; *p*=0.318). However, two-year hierarchal LME analyses revealed significant between-group differences. Compared to the no-NPS group, MBI-apathy status was associated with significantly higher p-tau_181_ levels over two years (26.46% [7.24% – 49.12%]; FDR-adjusted *p*=0.012), while non-MBI NPS status was not (11.18% [-2.84% – 27.23]; FDR-adjusted *p*=0.126). Tests of interactions between NPS group and time were not statistically significant (*p*>0.05), suggesting that, within this two-year period, there were no detectable differences in the slopes of change in p-tau_181_ between the groups.

Longitudinal biomarker data for plasma p-tau_181_ were available for 186/396 participants at the three-year mark (Table 4 and Figure 3).

**Table 4.** Longitudinal association between NPS group and plasma p-tau_181_ over three years modelled using linear mixed effect models adjusted for covariates.

| Predictors | Percentage Difference $\beta$ | 95% CI | $p$-value | FDR-adjusted $p$-value |
| --- | --- | --- | --- | --- |
| Non-MBI NPS | 10.78% | -2.81% – 26.27% | 0.128 | 0.128 |
| MBI-apathy | 29.28% | 10.17% – 51.72% | **0.002** | **0.004** |
| Time (Months) | 0.18% | -0.04% – 0.40% | 0.108 |  |
| Age | 1.94% | 1.17% – 2.71% | **<0.001** |  |
| Sex (Female) | -4.61% | -14.66% – 6.64% | 0.411 |  |
| Years of Education | 0.05% | -2.06% – 2.21% | 0.961 |  |
| APOE4 Carrier Status (APOE4 Carrier) | 22.58% | 9.52% – 37.19% | **<0.001** |  |
| MMSE Score | -2.69% | -6.04% – 0.78% | 0.130 |  |
| NPS status (Non-MBI NPS) × Time | -0.09% | -0.45% – 0.26% | 0.617 |  |
| NPS status (MBI-apathy) × Time | -0.09% | -0.51% – 0.33% | 0.680 |  |

Beta coefficients for the NPS group variables (highlighted in grey) represent the estimated percent difference in p-tau_181_ compared to the no-NPS group over three years. Tests of interaction between NPS group and time were non-significant. This model is adjusted for age, sex, years of education, APOE4 carrier status, MMSE score, and an interaction term between NPS status and number of months. APOE: Apolipoprotein E; CI: Confidence interval; FDR: False Discovery Rate; MBI: Mild Behavioural Impairment; MCI: Mild Cognitive Impairment; MMSE: Mini-Mental State Examination; NPS: Neuropsychiatric Symptoms.

Participants missing plasma p-tau_181_ data at three years were older (73.60 [6.70] *vs*. 71.25 [7.36] years, *p* = 0.001) and had higher MMSE scores (28.84 [1.45] *vs.* 28.15 [1.67], *p* < 0.001) compared to those with data available at that timepoint. Sex distribution (Female: 46.67% *vs.* 52.69%, *p* = 0.273), years of education (16.32 [2.64] *vs.* 16.34 [2.64], *p* = 0.925), and APOE4 carriership (35.71% *vs.* 43.55%, *p* = 0.136) did not significantly differ between groups. On average, plasma p-tau_181_ did not significantly change over three years across the full sample (0.18% [-0.04% – 0.40%]; *p*=0.108). However, three-year hierarchal LME analyses revealed that compared to the No-NPS group, MBI-apathy was associated with higher p-tau_181_ over three years (29.28% [10.17% – 51.72%]; FDR-adjusted *p* =0.004), while non-MBI NPS was not (10.78% [-2.81% – 26.27]; FDR-adjusted *p*=0.128). Tests of interactions between NPS group and time were not statistically significant (*p*>0.05), suggesting that, within the limits of our study, there are no detectable differences in the slopes of changes in p-tau_181_ between the groups.

## Discussion

In this study of 396 dementia-free older adults, we examined associations between MBI-apathy and blood plasma p-tau_181_ levels. Associations were assessed cross-sectionally and longitudinally over two and three years, comparing individuals with MBI-apathy and non-MBI NPS to those without NPS. Apathy symptoms meeting MBI criteria were consistently associated with higher plasma p-tau_181_ levels at baseline and follow-up, whereas non-MBI NPS showed no significant association.

Prior work from our group showed that, in CN and MCI individuals, MBI-apathy was associated with greater CSF tau pathology.^31^ In ADNI, individuals with MBI-apathy exhibited significantly higher CSF p-tau_181_ and CSF p-tau_181_/Aβ_42_ at baseline and over two years, while non-MBI NPS did not significantly differ from no-NPS. A four-year analysis in ADNI and MEMENTO cohorts supports this pattern at the global MBI level, although apathy was not examined specifically.^38^ Across four years, MBI was associated with higher CSF p-tau_181_ and CSF p-tau_181_/Aβ_42_, while non-MBI NPS showed no significant association. More recently, MBI has also been significantly associated with CSF Aβ_42_ and p-tau_181_ positivity, as well as with the NIA-AA defined AD continuum biomarker profile, whereas non-MBI NPS showed no associations.^39^ Concordant results were obtained with plasma markers for p-tau_181_ and p-tau_217_.^40, 41^ Global MBI was associated with significantly higher plasma p-tau_181_ at baseline and over four years, while non-MBI NPS again showed no difference.^40^ Global MBI was also significantly associated with higher plasma p-tau_217_ at baseline compared to those without MBI.^41^ Similarly, higher MBI Checklist scores have been associated with both elevated CSF p-tau_181_ and higher tau-PET signal in the entorhinal cortex and hippocampus among CN Aβ-positive individuals.^42^ Extending these findings, MBI status has been associated with greater tau tracer uptake in early AD cortical regions among Aβ-positive but not Aβ-negative individuals, reinforcing its role as a behavioural correlate of AD-related tau pathology.^43^ These biomarker results align with autopsy studies showing that MBI in the five years before death is significantly associated with pathologically confirmed AD, but not with non-AD neuropathologies, underscoring its specificity as a prodromal feature of AD.^44, 45^

Together, these findings show that applying the MBI framework to all later-life NPS helps identify the NPS subgroup most likely to have tau-related neurodegeneration. Specifically, MBI-apathy had the highest levels of p-tau_181_, helping identify preclinical and prodromal stages of the AD continuum. A key MBI feature is symptom persistence. Across all cohorts, elevated tau was observed only when symptoms had later-life onset and persisted for at least six months. In contrast, impersistent NPS, such as those captured in the non-MBI NPS group, can reflect situational, psychological, or comorbid medical factors unrelated to neurodegeneration.^18^ Pooling impersistent with persistent symptoms weakens observed biomarker associations, while excluding episodic cases sharpens the signal. In practice, applying MBI criteria does more than refine a case definition, it isolates a behavioural phenotype that more reliably indexes early AD pathology. Therefore, future biomarker studies and clinical trials should distinguish MBI from non-MBI NPS, as these groups differ in likelihood of underlying AD pathology. While most biomarker studies have examined MBI globally, apathy warrants particular attention as one of the most common and clinically consequential NPS.^46^ Our group has also specifically focused on the apathy domain, where associations with AD pathology have been consistent.^31^ MBI-apathy is therefore a particularly relevant domain for testing whether such relationships are also observable in plasma biomarkers and whether they persist over time.

Plasma p-tau_181_ levels remained higher in the MBI-apathy group over time; however, our longitudinal LMEs did not identify significant interaction effects between NPS group and time. As shown in Figure 3, the MBI-apathy group had roughly parallel trajectories to the other groups. The non-significant interaction suggests that, within the study timeframe, group trajectories in p-tau_181_ remained relatively stable and did not diverge significantly. The observed stability is consistent with AD biomarker staging models, which suggest that plasma p-tau_181_ rises after detectable amyloid accumulation and begins to plateau during late-MCI to early-dementia.^1, 5, 11, 37^ In contrast, longitudinal data from sporadic and autosomal dominant cohorts show that p-tau_217_ increases earlier in the AD cascade.^1, 4, 5, 11, 47^ As p-tau_181_ increases later in the disease course, it may better reflect downstream progression rather than initial onset. The absence of a significant interaction however, does not necessarily confirm identical slopes across groups. Absence of slope differences may indicate a biological inflection point occurring before our baseline. This inflection point coincides with global MBI, the construct for which was developed explicitly for early detection; MBI can initially manifest as the emergence and persistence of an undifferentiated constellation of symptoms prior to an identifiable MBI domain.^31, 40, 48^ Group-level differences in p-tau_181_ trajectories may therefore have already stabilized by enrolment, limiting our ability to detect divergence over the observed two- to three-year period. Short follow-up duration, modest assay sensitivity for detecting subtle change in plasma p-tau_181_, inter-individual variability, and attrition-related reductions in sample size could have further contributed to the absence of slope differences. Nonetheless, the consistently elevated p-tau_181_ levels observed in the MBI-apathy group, independent of trajectory acceleration, underscores its potential utility as a behavioural marker of early tau pathology.

The elevated risk profile of the MBI-apathy group is further underscored by its demographic characteristics. In our sample, those with MBI-apathy were more likely to carry the APOE4 allele, have MCI, and be male, features that, while adjusted for in all models, highlight clinical differences that may contribute to the early identification of at-risk individuals. These findings align with prior work from our group. For example, in a National Alzheimer’s Coordinating Center analysis, MBI-apathy cases were more often male (65.0%) and had higher MCI prevalence (58.5%) than those without apathy (39.7% male, 20.0% with MCI) and no NPS (37.4% male, 13.4% with MCI).^24^ Similarly, in our ADNI CSF biomarker study, more APOE4 carriers (59.6%) and males (67.3%) were observed among those with MBI-apathy compared to those with no NPS (48.8% male, 33.0% APOE4 carriers) and non-MBI NPS (45.7% male, 31.9% APOE4 carriers).^31^ Although male sex is not a risk factor for AD, and women have a higher lifetime risk,^49^ several studies suggest that apathy may be more common or severe in males.^19, 50, 51^ A meta-analysis showed that across AD clinical stages, apathy severity was higher in men, in contrast to depression or anxiety, which were more prevalent in women.^50^ Sex differences may reflect differential involvement of fronto-striatal circuits or dopaminergic tone, both of which are implicated in apathy.^52^ Differences in symptom reporting or recognition may also contribute, as men may be less likely to report affective complaints, instead presenting with motivational withdrawal. In addition, study partner characteristics may shape symptom detection and endorsement. Female study partners are more likely than male study partners to report NPS, which may influence observed sex differences.^53^ Taken together, these demographic patterns suggest that MBI-apathy clusters with other established indicators of elevated AD risk. Understanding the intersection of behavioural, genetic, and cognitive features could improve our ability to stratify risk and tailor early intervention strategies.

### Limitations

This study has a few relevant limitations that should be mentioned. First, the ADNI dataset is derived from a highly curated research cohort that was designed to be enriched for AD pathology. As such, participants are selected based on strict criteria, including minimal vascular burden and elevated risk of AD, making the cohort less representative of the general aging population. Additionally, the sample has limited ethnocultural and sociodemographic diversity, further restricting generalizability.

The MBI-apathy group is relatively small. While this was anticipated given the specificity of the MBI criteria, it limits our ability to examine more nuanced relationships. For instance, we were unable to assess interactions between NPS status and clinical cognitive diagnosis (*i.e.,* CN *vs.* MCI), as the number of CN individuals with MBI-apathy was too small for meaningful stratification (n = 5 at two years; n = 1 at three years).The limited sample also precluded analysis of plasma biomarkers beyond p-tau_181_, including p-tau_217_, despite growing evidence supporting p-tau_217_ as a more sensitive and specific marker of AD pathology than p-tau_181_. ^3, 11–17^ Indeed, our research group has recently found an association between MBI and plasma p-tau_217_,^41^ highlighting the need for future studies with larger samples and broader biomarker panels to clarify whether MBI-apathy shows similar associations.

In addition, apathy was operationalized using the NPI and NPI-Q, instruments originally developed for dementia populations. These tools may have limited sensitivity for detecting subtle or early motivational changes in non-dementia individuals, particularly when using the brief

NPI-Q, potentially underestimating apathy severity or prevalence in preclinical stages. Future studies using the MBI Checklist might be informative, as the MBI-C explicitly identifies apathy and the three apathy domains of interest, initiative, and emotional reactivity.^54–56^

Additionally, plasma p-tau_181_ was analyzed as a continuous outcome in this study, allowing detection of subtle group-level differences but not whether individuals with MBI-apathy were more likely to exceed clinical or pathological cutoffs. Future research incorporating binary classifications of biomarker positivity (*e.g.,* normal *vs.* abnormal plasma p-tau_181_) and comparison with non-MBI apathy could further clarify the prognostic relevance of persistent, later-life emergent apathy for risk stratification and early detection. Finally, our longitudinal analyses relied on LME models, which assume linear trajectories. Although diagnostic checks supported model adequacy, it is possible that non-linear changes in plasma p-tau_181_ were not fully captured. Future studies with larger samples and extended follow-up could examine flexible modeling approaches to better account for potential non-linearity.

### Conclusions

MBI-apathy represents a clinically meaningful behavioural syndrome that aligns with biological and cognitive markers of AD risk. In this study, emergent and persistent apathy symptoms, defined using the MBI framework, were associated with elevated plasma p-tau_181_, reinforcing the link between later-life behavioural change and early tau pathology. These findings build on a growing body of research supporting the utility of MBI for sample enrichment and risk stratification in preclinical and prodromal AD.

As blood-based biomarkers gain traction in both research and clinical settings, incorporating scalable behavioural indicators, such as MBI-apathy, offers a complementary and pragmatic approach to early detection. Relevance is heightened in contexts where biomarker access remains limited or cost-prohibitive. Screening for emergent and persistent apathy symptoms may help identify individuals who would benefit from closer monitoring, timely counselling, or preventative intervention. Future research should continue to explore how integrating behavioural and biological data can improve prognostic accuracy and inform targeted, stage-specific care strategies in AD.

## Data Availability

Redistribution of ADNI data is prohibited as per the ADNI Data Sharing Publication Policy and Data Use Agreement (https://adni.loni.usc.edu/wp-content/uploads/how_to_apply/ADNI_DSP_Policy.pdf). Data from ADNI are only available by request and can be found here: https://adni.loni.usc.edu/data-samples/access-data/#access_data. Authors can share data cleaning script with interested parties.

## Acknowledgements

Data used in preparation of this article were obtained from the ADNI database (https://adni.loni.usc.edu/). As such, the investigators within the ADNI contributed to the design and implementation of ADNI and/or provided data but did not participate in analysis or writing of this report. A complete listing of ADNI investigators can be found at: https://adni.loni.usc.edu/wp-content/uploads/how_to_apply/ADNI_Acknowledgement_List.pdf

## Author Contributions

Daniella Vellone (Conceptualization; Data curation; Formal analysis; Funding acquisition; Methodology; Project administration; Software; Visualization; Writing – original draft; Writing – review & editing); Rebeca Leon (Data curation; Methodology; Software; Writing – review & editing); Zahra Goodarzi (Supervision, Writing – review & editing); Nils D. Forkert (Writing – review & editing); Eric E. Smith (Writing – review & editing); Zahinoor Ismail (Conceptualization; Funding acquisition; Resources; Supervision, Writing – original draft; Writing – review & editing).

## Statements and Declarations

### Ethical Considerations

ADNI, which includes ADNI-1, ADNI-GO, ADNI-2, and ADNI-3, adheres to Good Clinical Practice guidelines, the ethical principles of the Declaration of Helsinki, US regulatory requirements (21 CFR Part 50: Protection of Human Subjects and Part 56: Institutional Review Boards), the Tri-Council Policy Statement: Ethical Conduct for Research Involving Humans (TCPS2), as well as Health Canada and International Committee on Harmonization of Good Clinical Practice guidelines. Study protocols were reviewed and approved by the Institutional Review Boards (IRBs) at all participating sites, including the IRB at the University of California, San Francisco, where the ADNI Coordinating Center is located. The full protocols and associated ethical procedures are publicly available through the ADNI website (https://adni.loni.usc.edu/help-faqs/adni-documentation/).

Participant privacy and data confidentiality were protected through the use of coded research identifiers; no personally identifying information was linked to the datasets used in this analysis. De-identified data were made publicly available through the ADNI data repository in line with the National Institute of Health data sharing policies.

This study involved a secondary analysis of existing de-identified ADNI data. As such, additional ethics approval was not required by the authors’ institution.

### Consent to Participate

All ADNI participants provided written informed consent prior to the initiation of any study procedures, including consent for data collection, storage, and future use in research. Informed consent included permission to collect and store biospecimens (e.g., blood and CSF), neuroimaging data, and longitudinal clinical assessments for research purposes. Where required, consent was also obtained from legally authorized representatives. Separate consent was obtained for genetic analyses and autopsy, in accordance with site-specific IRB requirements.

### Consent for Publication

Participants provided written informed consent authorizing the use of de-identified data and biospecimens for future research dissemination, including scientific publication. Consent documents specified that only coded, non-identifiable data would be shared and used in publications. No identifying personal information or images are reported in this manuscript.

### Data Availability

Redistribution of ADNI data is prohibited as per the ADNI Data Sharing Publication Policy and Data Use Agreement (https://adni.loni.usc.edu/wp-content/uploads/how_to_apply/ADNI_DSP_Policy.pdf). Data from ADNI are only available by request and can be found here: https://adni.loni.usc.edu/data-samples/access-data/#access_data.

Authors can share data cleaning script with interested parties.

## References

1. Jack CR, Jr., Andrews JS, Beach TG, et al. Revised criteria for diagnosis and staging of Alzheimer’s disease: Alzheimer’s Association Workgroup. Alzheimers Dement 2024 20240627. DOI: 10.1002/alz.13859.

2. Sperling R, Mormino E and Johnson K. The evolution of preclinical Alzheimer’s disease: implications for prevention trials. Neuron 2014; 84: 608–622. 20141105. DOI: 10.1016/j.neuron.2014.10.038.

3. Barthélemy NR, Li Y, Joseph-Mathurin N, et al. A soluble phosphorylated tau signature links tau, amyloid and the evolution of stages of dominantly inherited Alzheimer’s disease. Nat Med 2020; 26: 398–407. 20200311. DOI: 10.1038/s41591-020-0781-z.

4. Mattsson-Carlgren N, Andersson E, Janelidze S, et al. Aβ deposition is associated with increases in soluble and phosphorylated tau that precede a positive Tau PET in Alzheimer’s disease. Sci Adv 2020; 6: eaaz2387. 20200415. DOI: 10.1126/sciadv.aaz2387.

5. Therriault J, Vermeiren M, Servaes S, et al. Association of Phosphorylated Tau Biomarkers With Amyloid Positron Emission Tomography vs Tau Positron Emission Tomography. JAMA Neurol 2023; 80: 188–199. DOI: 10.1001/jamaneurol.2022.4485.

6. Fagan AM, Mintun MA, Mach RH, et al. Inverse relation between in vivo amyloid imaging load and cerebrospinal fluid Abeta42 in humans. Ann Neurol 2006; 59: 512–519. DOI: 10.1002/ana.20730.

7. de Wilde A, Reimand J, Teunissen CE, et al. Discordant amyloid-β PET and CSF biomarkers and its clinical consequences. Alzheimers Res Ther 2019; 11: 78. 20190912. DOI: 10.1186/s13195-019-0532-x.

8. Palmqvist S, Mattsson N and Hansson O. Cerebrospinal fluid analysis detects cerebral amyloid-β accumulation earlier than positron emission tomography. Brain 2016; 139: 1226–1236. 20160302. DOI: 10.1093/brain/aww015.

9. Rajmohan R and Reddy PH. Amyloid-Beta and Phosphorylated Tau Accumulations Cause Abnormalities at Synapses of Alzheimer’s disease Neurons. J Alzheimers Dis 2017; 57: 975–999. DOI: 10.3233/jad-160612.

10. Suárez-Calvet M, Karikari TK, Ashton NJ, et al. Novel tau biomarkers phosphorylated at T181, T217 or T231 rise in the initial stages of the preclinical Alzheimer’s continuum when only subtle changes in Aβ pathology are detected. EMBO Mol Med 2020; 12: e12921. 20201110. DOI: 10.15252/emmm.202012921.

11. Teunissen CE, Kolster R, Triana-Baltzer G, et al. Plasma p-tau immunoassays in clinical research for Alzheimer’s disease. Alzheimers Dement 2025; 21: e14397. 20241203. DOI: 10.1002/alz.14397.

12. Mattsson-Carlgren N, Janelidze S, Palmqvist S, et al. Longitudinal plasma p-tau217 is increased in early stages of Alzheimer’s disease. Brain 2020; 143: 3234–3241. DOI: 10.1093/brain/awaa286.

13. Palmqvist S, Warmenhoven N, Anastasi F, et al. Plasma phospho-tau217 for Alzheimer’s disease diagnosis in primary and secondary care using a fully automated platform. Nat Med 2025 20250409. DOI: 10.1038/s41591-025-03622-w.

14. Mendes AJ, Ribaldi F, Lathuiliere A, et al. Head-to-head study of diagnostic accuracy of plasma and cerebrospinal fluid p-tau217 versus p-tau181 and p-tau231 in a memory clinic cohort. J Neurol 2024; 271: 2053–2066. 20240109. DOI: 10.1007/s00415-023-12148-5.

15. Kwon HS, Hwang M, Koh SH, et al. Comparison of plasma p-tau217 and p-tau181 in predicting amyloid positivity and prognosis among Korean memory clinic patients. Sci Rep 2025; 15: 7791. 20250306. DOI: 10.1038/s41598-025-90232-8.

16. Pandey N, Yang Z, Cieza B, et al. Plasma phospho-tau217 as a predictive biomarker for Alzheimer’s disease in a large south American cohort. Alzheimers Res Ther 2025; 17: 1. 20250102. DOI: 10.1186/s13195-024-01655-w.

17. Lin YS, Kwon HS, Lee WJ, et al. Cross-cultural validation of plasma p-tau217 and p-tau181 as precision biomarkers for amyloid PET positivity: An East Asian study in Taiwan and Korea. Alzheimers Dement 2025; 21: e14565. DOI: 10.1002/alz.14565.

18. Ismail Z, Smith EE, Geda Y, et al. Neuropsychiatric symptoms as early manifestations of emergent dementia: Provisional diagnostic criteria for mild behavioral impairment. Alzheimers Dement 2016; 12: 195–202. 20150618. DOI: 10.1016/j.jalz.2015.05.017.

19. Wolfova K, Creese B, Aarsland D, et al. Gender/Sex Differences in the Association of Mild Behavioral Impairment with Cognitive Aging. J Alzheimers Dis 2022; 88: 345–355. DOI: 10.3233/jad-220040.

20. Rouse HJ, Ismail Z, Andel R, et al. Impact of Mild Behavioral Impairment on Longitudinal Changes in Cognition. J Gerontol A Biol Sci Med Sci 2024; 79. DOI: 10.1093/gerona/glad098.

21. Ghahremani M, Smith EE and Ismail Z. Improving dementia prognostication in cognitively normal older adults: conventional versus novel approaches to modelling risk associated with neuropsychiatric symptoms. The British Journal of Psychiatry 2025; 226: 129–136. 2024/12/16. DOI: 10.1192/bjp.2024.136.

22. Kan CN, Cano J, Zhao X, et al. Prevalence, Clinical Correlates, Cognitive Trajectories, and Dementia Risk Associated With Mild Behavioral Impairment in Asians. J Clin Psychiatry 2022; 83 20220316. DOI: 10.4088/JCP.21m14105.

23. Ebrahim IM, Ghahremani M, Camicioli R, et al. Effects of race, baseline cognition, and APOE on the association of affective dysregulation with incident dementia: A longitudinal study of dementia-free older adults. J Affect Disord 2023; 332: 9–18. 20230328. DOI: 10.1016/j.jad.2023.03.074.

24. Vellone D, Ghahremani M, Goodarzi Z, et al. Apathy and APOE in mild behavioral impairment, and risk for incident dementia. Alzheimer’s Dement: Transl Res Clin Interv 2022; 8: 1–12. DOI: 10.1002/trc2.12370.

25. Zahinoor Ismail, Maryam Ghahremani, Amlish M. Munir, et al. A longitudinal study of late-life psychosis and incident dementia and the potential effects of race and cognition. Nature Mental Health 2023; 1: 273–283.

26. Creese B, Arathimos R, Aarsland D, et al. Late-life onset psychotic symptoms and incident cognitive impairment in people without dementia: Modification by genetic risk for Alzheimer’s disease. Alzheimers Dement (N Y) 2023; 9: e12386. 20230430. DOI: 10.1002/trc2.12386.

27. Marin RS. Differential diagnosis and classification of apathy. Am J Psychiatry 1990; 147: 22–30. DOI: 10.1176/ajp.147.1.22.

28. Marin RS. Apathy: a neuropsychiatric syndrome. J Neuropsychiatry Clin Neurosci 1991; 3: 243–254. DOI: 10.1176/jnp.3.3.243.

29. Miller DS, Robert P, Ereshefsky L, et al. Diagnostic criteria for apathy in neurocognitive disorders. Alzheimers Dement 2021; 17: 1892–1904. 20210505. DOI: 10.1002/alz.12358.

30. Chong TT. Definition: Apathy. Cortex 2020; 128: 326–327. 20200410. DOI: 10.1016/j.cortex.2020.04.001.

31. Vellone D, Leon R, Goodarzi Z, et al. Mild behavioural impairment-apathy and core Alzheimer’s disease cerebrospinal fluid biomarkers. Brain 2025 20250602. DOI: 10.1093/brain/awaf194.

32. Kang MJY, Eratne D, Loi SM, et al. Apathy and affective symptoms associated with elevated plasma neurofilament light but not p-tau181 in Alzheimer’s disease. Alzheimers Dement (Amst) 2025; 17: e70151. 20250729. DOI: 10.1002/dad2.70151.

33. Hall JR, Petersen M, Johnson L, et al. Plasma Total Tau and Neurobehavioral Symptoms of Cognitive Decline in Cognitively Normal Older Adults. Front Psychol 2021; 12: 774049. 20211105. DOI: 10.3389/fpsyg.2021.774049.

34. Cummings JL, Mega M, Gray K, et al. The Neuropsychiatric Inventory: comprehensive assessment of psychopathology in dementia. Neurology 1994; 44: 2308–2314. DOI: 10.1212/wnl.44.12.2308.

35. Cummings J. The Neuropsychiatric Inventory: Development and Applications. J Geriatr Psychiatry Neurol 2020; 33: 73–84. DOI: 10.1177/0891988719882102.

36. Guan DX, Smith EE, Pike BG, et al. Persistence of neuropsychiatric symptoms and dementia prognostication: A comparison of three operational case definitions of mild behavioral impairment. Alzheimers Dementia 2023; 15: e12483.

37. Karikari TK, Pascoal TA, Ashton NJ, et al. Blood phosphorylated tau 181 as a biomarker for Alzheimer’s disease: a diagnostic performance and prediction modelling study using data from four prospective cohorts. Lancet Neurol 2020; 19: 422–433. DOI: 10.1016/s1474-4422(20)30071-5.

38. Ismail Z, Leon R, Creese B, et al. Optimizing detection of Alzheimer’s disease in mild cognitive impairment: a 4-year biomarker study of mild behavioral impairment in ADNI and MEMENTO. Mol Neurodegener 2023; 18: 50. 20230729. DOI: 10.1186/s13024-023-00631-6.

39. Leon R, Ghahremani M, Guan DX, et al. Enhancing Alzheimer Disease Detection Using Neuropsychiatric Symptoms: The Role of Mild Behavioural Impairment in the Revised NIA-AA Research Framework. J Geriatr Psychiatry Neurol 2025: 8919887251366634. 20250813. DOI: 10.1177/08919887251366634.

40. Ghahremani M, Wang M, Chen HY, et al. Plasma Phosphorylated Tau at Threonine 181 and Neuropsychiatric Symptoms in Preclinical and Prodromal Alzheimer Disease. Neurology 2023; 100: e683–e693. 20221102. DOI: 10.1212/wnl.0000000000201517.

41. Ghahremani M, Leon R, Smith EE, et al. Exploring the association between mild behavioral impairment and plasma p-tau217: Implications for early detection of Alzheimer’s disease. Alzheimers Dement (Amst) 2025; 17: e70119. 20250521. DOI: 10.1002/dad2.70119.

42. Johansson M, Stomrud E, Insel PS, et al. Mild behavioral impairment and its relation to tau pathology in preclinical Alzheimer’s disease. Transl Psychiatry 2021; 11: 76. 20210126. DOI: 10.1038/s41398-021-01206-z.

43. Naude J, Wang M, Leon R, et al. Tau-PET in early cortical Alzheimer brain regions in relation to mild behavioral impairment in older adults with either normal cognition or mild cognitive impairment. Neurobiol Aging 2024; 138: 19–27. 20240216. DOI: 10.1016/j.neurobiolaging.2024.02.006.

44. Sharif SF, Guan DX, Bodnar T, et al. Neuropsychiatric symptoms and progression to pathologically confirmed Alzheimer’s disease. Brain 2025 20250425. DOI: 10.1093/brain/awaf156.

45. Ruthirakuhan M, Ismail Z, Herrmann N, et al. Mild behavioral impairment is associated with progression to Alzheimer’s disease: A clinicopathological study. Alzheimer’s & Dementia 2022; 18: 2199–2208. DOI: 10.1002/alz.12519.

46. Mortby ME, Adler L, Agüera-Ortiz L, et al. Apathy as a Treatment Target in Alzheimer’s Disease: Implications for Clinical Trials. Am J Geriatr Psychiatry 2022; 30: 119–147. 20210701. DOI: 10.1016/j.jagp.2021.06.016.

47. McDade E, Wang G, Gordon BA, et al. Longitudinal cognitive and biomarker changes in dominantly inherited Alzheimer disease. Neurology 2018; 91: e1295–e1306. 20180914. DOI: 10.1212/WNL.0000000000006277.

48. Miao R, Chen HY, Robert P, et al. White matter hyperintensities and mild behavioral impairment: Findings from the MEMENTO cohort study. Cereb Circ Cogn Behav 2021; 2: 100028. 20210914. DOI: 10.1016/j.cccb.2021.100028.

49. 2024 Alzheimer’s disease facts and figures. Alzheimers Dement 2024; 20: 3708–3821. 20240430. DOI: 10.1002/alz.13809.

50. Eikelboom WS, Pan M, Ossenkoppele R, et al. Sex differences in neuropsychiatric symptoms in Alzheimer’s disease dementia: a meta-analysis. Alzheimers Res Ther 2022; 14: 48. 20220404. DOI: 10.1186/s13195-022-00991-z.

51. Guan DX, Chen HY, Camicioli R, et al. Dual-task gait and mild behavioral impairment: The interface between non-cognitive dementia markers. Exp Gerontol 2022; 162: 111743. 20220216. DOI: 10.1016/j.exger.2022.111743.

52. Zachry JE, Nolan SO, Brady LJ, et al. Sex differences in dopamine release regulation in the striatum. Neuropsychopharmacology 2021; 46: 491–499. 20201214. DOI: 10.1038/s41386-020-00915-1.

53. Guan DX, Mudalige D, Munro CE, et al. The effect of study partner characteristics on the reporting of neuropsychiatric symptoms across the neurocognitive spectrum. Int Psychogeriatr 2024; 36: 675–688. 20240918. DOI: 10.1017/s1041610224000590.

54. Hu S, Patten S, Charlton A, et al. Validating the Mild Behavioral Impairment Checklist in a Cognitive Clinic: Comparisons With the Neuropsychiatric Inventory Questionnaire. J Geriatr Psychiatry Neurol 2022: 8919887221093353. 20220417. DOI: 10.1177/08919887221093353.

55. Ismail Z, Aguera-Ortiz L, Brodaty H, et al. The Mild Behavioral Impairment Checklist (MBI-C): A Rating Scale for Neuropsychiatric Symptoms in Pre-Dementia Populations. J Alzheimers Dis 2017; 56: 929–938. DOI: 10.3233/JAD-160979.

56. Vellone DA, Guan DX and Ismail Z. The Association Between Mild Behavioral Impairment-Apathy Symptoms and Cognitive Symptoms. Alzheimers Dement 2024; 20 20250103. DOI: 10.1002/alz.090525.

